# Potential biomarkers for fatal outcome prognosis in a cohort of hospitalized COVID-19 patients with pre-existing co-morbidities

**DOI:** 10.1101/2023.04.25.23288937

**Authors:** Ruth Lizzeth Madera-Sandoval, Arturo Cérbulo-Vázquez, Lourdes Andrea Arriaga-Pizano, Graciela Libier Cabrera-Rivera, Edna Basilio-Gálvez, Patricia Esther Miranda-Cruz, María Teresa García de la Rosa, Jessica Lashkmin Prieto-Chávez, Silvia Vanessa Rivero-Arredondo, Alonso Cruz-Cruz, Daniela Rodríguez-Hernández, María Eugenia Salazar-Ríos, Enrique Salazar-Ríos, Esli David Serrano-Molina, Roberto Carlos De Lira-Barraza, Abel Humberto Villanueva-Compean, Alejandra Esquivel-Pineda, Rubén Ramírez-Montes de Oca, Omar Unzueta-Marta, Guillermo Flores-Padilla, Juan Carlos Anda-Garay, Luis Alejandro Sánchez-Hurtado, Salvador Calleja-Alarcón, Laura Romero-Gutiérrez, Rafel Torres-Rosas, Laura C. Bonifaz, Rosana Pelayo, Edna Márquez-Márquez, Constantino III Roberto López-Macías, Eduardo Ferat-Osorio

## Abstract

**Background:** The difficulty to predict fatal outcomes in COVID-19 patients, impacts in the general morbidity and mortality due to SARSCoV2 infection, as it wears out the hospital services that care for these patients. Unfortunately, in several of the candidates for prognostic biomarkers proposed, the predictive power is compromised when patients have pre-existing co-morbidities.

**Methods:** A cohort of one hundred and forty-seven patients hospitalized for severe COVID19 was included in a descriptive, observational, single-center, and prospective study. Patients were recruited during the first COVID-19 pandemic wave (April-Nov, 2020). Data were collected from the clinical history while immunophenotyping by multiparameter flow cytometry analysis allowed us to assess the expression of surface markers on peripheral leukocytes. Patients were grouped according to the outcome in survivor or decease. The prognostic value of leukocytes, cytokines or HLA-DR, CD39, and CD73 was calculated.

**Results:** Hypertension and chronic renal failure but not obesity and diabetes were conditions more frequent among the decease group. Mixed hypercitokinemia, including inflammatory (IL-6) and anti-inflammatory (IL-10) cytokines, was more evident in deceased patients. In the decease group, lymphopenia with a higher NLR value was present. HLA-DR expression and the percentage of CD39+ cells were higher than non COVID-19 patients, but remain similar despite outcome. ROC analysis and cut-off value of NLR (69.6%, 9.4), pNLR (71.1%, 13.6), IL-6 (79.7%, 135.2 pg/mL).

**Conclusion:** The expression of HLA-DR, CD39, and CD73, as many serum cytokines (other than IL-6) and chemokines levels do not show prognostic potential compared to NLR and pNLR values.

## Introduction

The SARS-CoV-2 acute infection may evoke a strong inflammatory response [1]. Even a similitude with the Macrophage Activation Syndrome (MAS) or secondary Haemophagocytic Lymphohistocytosis (sHLH) was proposed early during the first COVID-19 pandemic wave [2]. In hospitalized patients with COVID19 an Adult Respiratory Distress Syndrome (ARDS) has arised as a common complication [3]. ARDS is associated with hyperinflammatory response. Certain co-morbidities with a proposed inflammatory landscape, such as hypertension, cardiovascular disease, diabetes, and obesity, are also associated with severity and high-risk mortality in COVID-19 [4, 5].

So hyperinflammation seems to be a typical component of severe or critic COVID-19 [6]. Inflammation is a complex response in patients with severe COVID-19 and may result in a clinical picture resembling sepsis [1, 7]. Like in sepsis, in COVID-19 some clinical characteristics and hyperinflammatory response could be used at admission as predictors of in-hospital death [4, 8, 9]. Cytokines such as IL-6 and IL-10 in serum could reach a high concentration in the serum of COVID-19 patients and have been explored as predictors of fatal outcomes [10-14]. Routine laboratory inflammatory markers such as NLR(Neutrophil Lymphocyte Ratio) may also predict a fatal outcome in these patients [15-19]. However, these predictors may be less accurate if patients have pre-existing co-morbidities. The difficulty to predict fatal outcomes in COVID-19 patients impacts in the general morbidity and mortality due to SARSCoV2 infection, as it wears out the hospital services that care for these patients.

Some surface molecules related with leukocyte activation during systemic inflammatory response (infectious and non-infectious) may be more stable potential prognostic biomarkers. HLA-DR diminished expression in monocytes or dendritic cells is associated with poor prognosis in patients with sepsis or acute pancreatitis [20-22]. Also, HLA-DR diminished expression has been reported in myeloid-derived suppressor cells from patients with COVID-19 [23], as well as in monocytes (CD14+cells) from critically ill patients [24]. Since COVID-19 evokes both inflammatory and anti-inflammatory response, CD39/CD73 on leucocytes could also help to monitorized the patients with COVID-19[25, 26]. CD39 and CD73 are surface enzymes on monocytes, T, and B cells that could elicit an anti-inflammatory [27, 28] or even immunosuppressive response [29]. This coexistence of multiple mediators with antagonistic functions could explain why functional capacities can be compromised and result in more tortuous clinical evolutions. Indeed, in COVID-19 patients CD8+ T cell population undergoes quantitative and qualitative changes in COVID-19 patients that are both associated with suboptimal responses and higher disease severity [30].

To determine, even in patients with pre-existing co-morbidities, whether the differential expression of HLA-DR, CD39, and CD73 on the surface of circulating leukocytes might represent early biomarkers of COVID-19 outcome, we analyzed the immunophenotype of circulating leukocytes in patients hospitalized for severe COVID-19 during the first COVID-19 pandemic wave. Other inflammatory markers such as cytokines, chemokines, and serum D-dimer levels, as well as white blood cell count and NLR (Neutrophil/Lymphocyte ratio) or percentage NLR (pNLR), were also explored as indicators of inflammation and potential biomarkers.

## Material and methods

### Patients

One hundred and forty-seven patients hospitalized for COVID19 at (hidden peer review process) from April 17^th^ 2020 to November 8^th^ 2020 were enrolled for a single center, prospective, observational, longitudinal study approved by the National Research and Ethics Committee (hidden peer review process).

SARS-CoV-2 viral infection was confirmed by specific reverse transcription– polymerase chain reaction (RT-PCR). The clinical signs, symptoms, and co-morbidities were registered for every COVID-19 patient, and the outcome was recorded as survivor or fatal. Our study is in accordance with the World Medical Association’s Declaration of Helsinki. After a patient agreed to participate in the study, healthcare personal collected blood specimens in silicone-coated heparinized tubes (BD Vacutainer, N.J, USA) and samples were processed immediately after their collection.

Also, a group of healthy donors was included (n=11); they were matched by age and co-morbidities with the COVID-19 patients and did not show any sign of COVID-19 at the moment of blood sample collection.

### Clinical evaluation

Demographic and clinical data were collected at admission. Medical history includes co-morbidity and its medical treatment. Clinical data includes symptoms and determinations of respiratory rate, heart rate, temperature, and arterial pressure. Patients were followed-up since admission and until hospital discharge, then patients were assigned to survival or decease group.

### Laboratory data

Routine laboratory test was performed (blood count, coagulation profile, serum biochemical test, fibrinogen, and D-dimer), and processed in the central laboratory and by flow cytometry center, colorimetry, chemiluminescence, and immunoturbidimetry, respectively.

### Serum cytokine and chemokine assessment

The plasma concentrations of cytokines (IL-1β, IL-6, TNF-α, IFN-λ-1, IL-12p70, IFN-α2, IFN-λ2/3, GM-CSF, IFN-β e IFN-γ) and chemokines (CXCL8/IL-8, CXCL10/IP-10, CCL11/Eotaxin, CCL17/TARC, CCL2/MCP-1, CCL5/RANTES, CCL3/MIP-1a, CXCL9/MIG, CXCL5/ENA-78, CCL20/ MIP-3a, CXCL1/GROa, CXCL11/I-TAC and CCL4/MIP-1b) were determined using bead based immunoassays (LEGENDplex, Cat. 740350, BioLegend, San Diego, CA, USA and LEGENDplex, Cat. 740003, BioLegend, San Diego, CA, USA), according to the manufacturer’s instructions. Log-transformed data were used to construct standard curves fitted to 10 discrete points using a 4-parameter logistic model. The concentrations in the test samples were calculated using interpolations of their corresponding standard curves.

### Leucocyte immunophenotyping

Whole blood samples (50 μL) were incubated with titrated volumes of antibodies according to the following panel: anti-CD45-PerCP (Clone: HI30), anti-CD3-APC/Cy7 (Clone: UCHT1), anti-CD14-PECy7 (Clone:M5E2), anti-CD16-FITC (Clone:3G8), anti-CD73-PE (Clone:AD2), anti-CD39-BV421 (Clone: A1) and anti-HLA-DR-PE/Dazzle594 (Clone: L243). After 15 minutes of incubation, erythrolysis was performed using FACS™ Lysing Solution (Cat. 349202, BD, San Jose, CA, USA). Samples were washed once with 1x PBS (1,500 rpm, 5 minutes, room temperature) and resuspended in PBS (100 μL). At least 30,000 leucocytes (CD45+ cells) were acquired in a FACSAria IIu flow cytometer (BD Biosciences, San José, CA, USA). The FACS files were analyzed with Infinicyt™ software 1.8 (Cytognos, Salamanca, Spain), and FlowJo v10 software (Becton Dickinson, New Jersey, USA). Single cells were defined with an FSC-A vs. FSC-H plot, and leucocytes were identified using an SSC vs. CD45 plot. Lymphocytes were gated as SSC^low^FSC^low^CD45^++^, monocytes as SSC^mid^FSC^mid^CD45^+^CD14^+/-^CD16^+/-^, and neutrophils as SSC^mid^FSC^mid^CD45^+^CD16^+^. Lymphocyte subtypes were identified according to CD3-HLA-DR+ (B-like cells) and CD3+ (T cells). Monocytes subpopulations were identified as classical CD14^++^ CD16^-^, intermediate as CD14^+^CD16^+^ and no classical as CD14^-^CD16^+^. The percentages and mean fluorescence intensities (MFIs) of HLA-DR, CD39, and CD73 were calculated.

### Statistical Analysis

Statistical analysis was performed using Graph Pad Prism® version 6 software (GraphPad Software, San Diego, CA, USA), the IBM SPSS Statistics version 25.0 (IBM, Armonk, NY, USA) and the R i386 3.5.2 terminal (Microsoft corp., Boston, MA, USA.). Normality statistic tests were run for every variable (Shapiro-Wilk test). Non-parametric ANOVA test (Kruskall-Wallis test) with Dunn post-test were applied. Also, categorical variables were expressed as number (%) and compared by Fisher’s exact test. To biomarker prognostic value and the optimal cut-off, we used the C-statics of receiver operator characteristic (ROC) and the optimal cut-off was estimated in accordance with Youden Index (J). Multivariate logistical regression model was performed for corroborate the independent predictive analysis, age, and IMC were included, Hosmer and Lemeshow test was used to check model adequacy for goodness-of-fit statistics. Kaplan-Meier survival analysis were used to compare the outcomes and Log-Rank test was applied to validated Hazard ratio (HR) and 95% confidential interval (95% CI). A *P < 0.05* value was considered statistically significative admitted.

## Results

A hundred and forty-seven patients were included in our study. The outcome was recorded, and patients were analyzed as survive (n=78) or decease group (n=69). Table 1 shows clinical and laboratory characteristics in COVID-19 patients, the mean age in the decease group was higher (59±14 years, mean±SD) than in survive group (49±14 years, mean±SD, p<0.0001). No statistical difference was observed between survive and decease groups for sex, body mass index, respiratory rate, heart rate, and temperature (Table 1). Also, we did not observe a statistical difference between the survive and decease groups for some co-morbidities like diabetes mellitus or obesity. However, we found a statistical difference in the frequency of hypertension (p=0.011) and IRC (p=0.016). The sequential organ failure assessment (SOFA) score is higher in the decease group than in survive patients and reaches high significance (p=0.0001). On the other hand, we observed some laboratory parameters with no statistical difference like in glucose, creatinine, ferritin, TPT, D-Dimer, fibrinogen, and clinical scores like PLR o PIIV (Table 1). The laboratory parameters that showed statistical differences between survive and decease groups such as leucocyte and lymphocyte count, NLR, DHL, TP, PCR, PCT, SAT O_2,_ and the clinical score SIRI (Table 1).

**Table 1.** Clinical and laboratory characteristics in COVID-19 patients.

|  | **Total**<br>**(n=147)** | **Survive**<br>**(n=78)a** | **Decease**<br>**(n=69)b** | ***p*** |
| --- | --- | --- | --- | --- |
| **Age (years)** | 53±18 | 49±14 | 59±14 | **<0.0001** |
| **Male/Female** | 100/45 | 56/21 | 44/24 | 0.297 |
| **BMI** | 29.8±6.1 | 29.0±4.9 | 30.7±7.3 | 0.114 |
| **Respiratory Rate**<br>**(breaths per min)** | 24.8±8.1 | 23.9±6.3 | 25.5±5.8 | 0.141 |
| **Heart Rate**<br>**(beats per min)** | 101±19.5 | 103.6±20.5 | 101.2±17.3 | 0.494 |
| **Temperature (°C)** | 36.7±0.5 | 36.5±0.9 | 36.6±0.6 | 0.524 |
| **Diabetes Mellitus (pos/neg)** | 54/92 | 24/54 | 30/38 | 0.095 |
| **Obesity (pos/neg)** | 59/81 | 28/49 | 31/32 | 0.125 |
| **Hypertension (pos/neg)** | 49/97 | 19/59 | 30/38 | **0.011** |
| **CRF (pos/neg)** | 16/129 | 4/73 | 12/56 | **0.016** |
| **Leucocytes** | 9.9±4.6 | 9.1±4.0 | 10.8±4.7 | **0.022** |
| **Lymphocytes** | 0.9±0.5 | 0.9±0.4 | 0.7±0.5 | **0.038** |
| **NLR** | 15.1±14.1 | 10.8±11.2 | 17.0±15.5 | **0.006** |
| **Glucose** | 149.9±80 | 143.5±87 | 157.1±87 | 0.345 |
| **Ferritin** | 1543±1248 | 1455±1285 | 1644±2495 | 0.590 |
| **Creatinine** | 2.1±2.1 | 1.4±3.5 | 2.8±5.6 | 0.059 |
| **LDH** | 538±211 | 436.5±162 | 647±537 | **0.001** |
| **TP** | 16±15.6 | 14.5±1.3 | 15.6±2.3 | **0.0008** |
| **TPT** | 31.3±16.2 | 31.6±17.3 | 35±33.8 | 0.458 |
| **PCR** | 17.3±10.3 | 14.8±9.4 | 19.8±10.3 | **0.004** |
| **PCT** | 2.9±3.3 | 1.1±2.8 | 4.8±11.5 | **0.021** |
| **D-Dimer** | 4.4±3.6 | 3.5±9.2 | 6.8±24.9 | 0.281 |
| **Fibrinogen** | 698±200 | 716±172 | 677±179 | 0.188 |
| **Sat O2** | 88.7±7.4 | 90.4±7.7 | 86.7±10.6 | **0.028** |
| **SOFA** | 3.8±2.8 | 2.8±1.9 | 5±3.4 | **0.0001** |
| **SIRI** | 6.3±9.6 | 4.4±7.4 | 8.3±12.2 | **0.021** |
| **PLR** | 402±234 | 371.4±216.8 | 438.5±261.2 | 0.091 |
| **PIIV** | 1781±3083 | 1474±2446 | 2139±2814 | 0.129 |

mean±SD. BMI; Body Mass Index. Kruskal-Wallis test. CRF; Chronic Renal Failure. NLR; Neutrophil Lymphocyte Ratio. LDH; TP; TPT; PCR; PCT; Significant $p<0.05$ .

The most common SARS-CoV-2-related symptoms were cough, dyspnea, myalgia, and arthralgia, in both survive and decease patients, without any statistical significance (Table 1). Also, plasma cytokines and chemokines were assessed and shown in Figure 1. IL-6, CXCL8, IL-10, CCL2, CCL4, CCL20, CXCL10, and CXCL11 levels were significantly higher in deceased patients than in healthy donors. Also, IL-6, IL-10, CXCL10, and CXCL11 is higher in survivor patient than in healthy donor (Fig 1). However, cytokines/chemokines plasma concentrations did not show significant differences between survive and decease patients (Fig S1).

**Fig 1.**
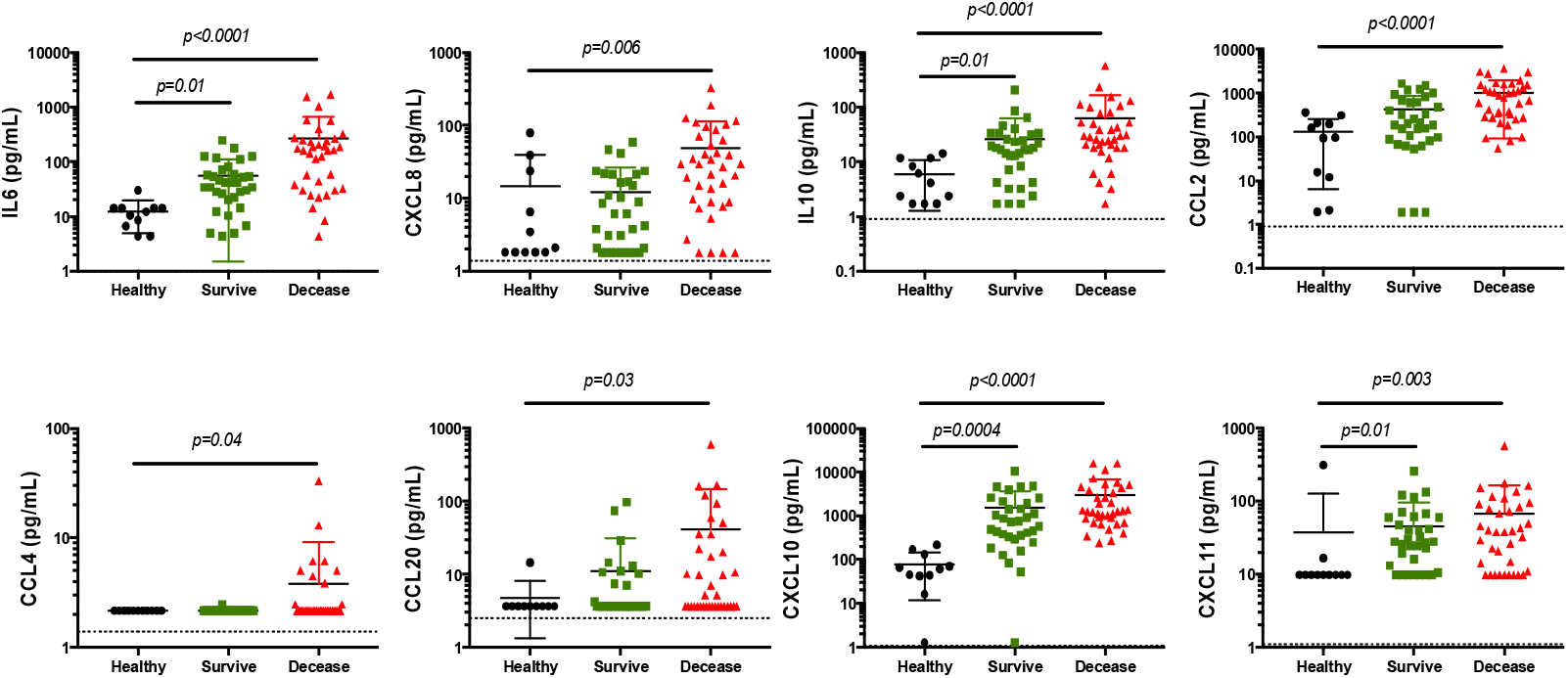
Plasma cytokine/chemokine concentration in COVID-19 patients. Cytokine and chemokines were assessed as in methods. Black circles: Healthy controls. Green squares: COVID-19 survive patients. Red triangles: COVID-19 fatal outcome patients. Dash line show the low limit of detection for each analyte. Results are expressed as mean±SD. Kruskal-Wallis and Dunn’s multiple comparisons test was calculated. Significant *p*<0.05.

Based on the differences we showed in Table 1 for NLR and pNLR, a ROC curve analysis was performed and showed for them a good prognostic values for clinical outcome. In the case of the NLR with an AUC of 0.700 (95% CI 0.624-0.777, p <0.0001). The cut-off for NLR was 9.4 with 66.0% and 64.9% of sensitivity and specificity respectively (Fig. 2c), and Kaplan-Meier curves show it as a good predictor for values >9.4 (p=0.0008, HR:43, 95%CI:0.261-0.685, Fig. 2d). The best discriminant of clinical outcome was pNLR with an AUC of 0.711 (95% CI 0.635-0.786, p <0.0001), and the optimal cut-off value was 13.6% with a sensitivity of 50.7% and specificity of 80.0% (Fig. 2e, Kaplan-Meier curves results were significant as a predictor for values >13.6% (p=0.011, HR:55, 95%CI:0.312-0.843, Fig. 2f). The ROC curve analysis for some cytokine/chemokine is shown in Fig. S2, where IL-6 has a good AUC 0.797 (95% CI 0.580-0.870, p= 0.005). The optimal cut-off value for IL-6 to predict mortality was 135.2 pg/mL (Fig. 2a) with a sensitivity of 61.5% and a specificity of 95.2%. Kaplan-Meier survival curves were performed, showing that IL-6 levels >135.2 pg/ml is a significant predictor for fatal outcome (Fig. 2b), (p=0.005, HR:43, 95%CI:0.283-0.786). A multivariate logistical regression model identifies NLR, pNLR, and IL6 as a good predictor, not the same for age and IMC (data do not show). Hosmer and Lemeshow test showed a good model adequacy (*X*^*2*^ = 4.6, p= 0.799).

**Fig 2.**
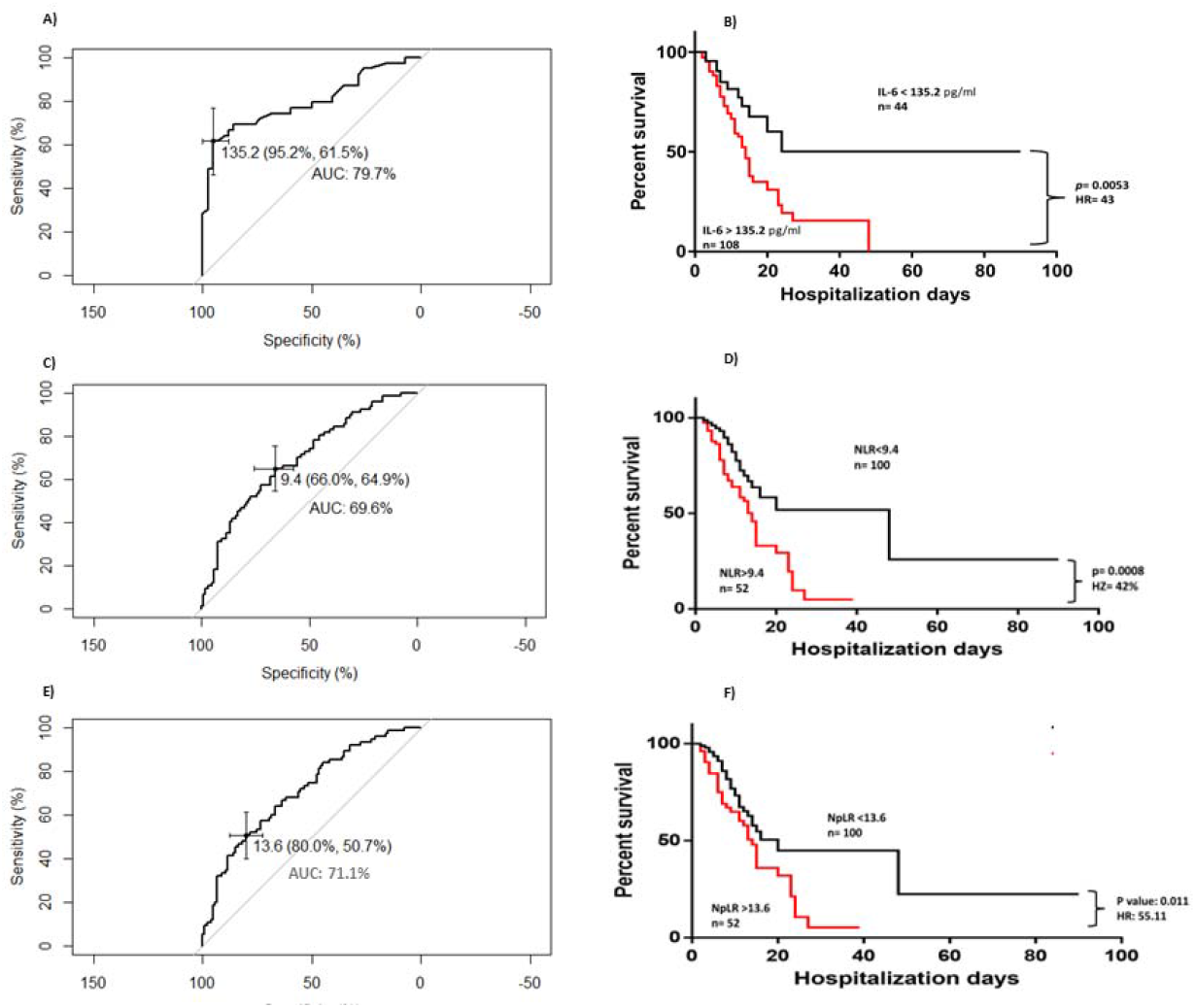
Receiver operator characteristics and survival curves for IL-6, NLR and pNLR in COVID-19 patients. Two different ROC curves were performed for IL-6 (A, B), NLR (C, D), and pNLR (E, F), optimal cut-off was calculated in accordance with Youden index. Kaplan-Meier of survival curves were made according to the optimal cut-off and HR expressed in percentage. Significant *p*<0.05, HR: Hazard ratio

About activation markers as HLA-DR was analyzed on the surface of leucocytes in survived and decease COVID-19 patients. Also, Figure 3 shows the percentage of HLA-DR+ on monocytes and T lymphocytes in healthy donors. Healthy donors express a high percentage of monocytes HLA-DR+, usually around 95% of monocytes, while the percentage of monocytes HLA-DR+ in COVID-19 patients is lower than 90 %. Figure 3A shows that on admission day (d0), the percentage of HLA-DR+ monocytes were significantly lower than in healthy donors, also a lower percentage were detected in COVID-19 patients after seventh days of medical treatment in survive (d7S) or decease (d7D) patients. Also, Fig 3A shows that the percentage of HLA-DR+ monocytes are higher in d7S than in d7D (Mann-Whitney test, 95% IC, p<0.0001). In addition, Fig 3 B, C, and D show the percentage of HLA-DR+ cells in Classical (Monocytes C),

**Fig 3.**
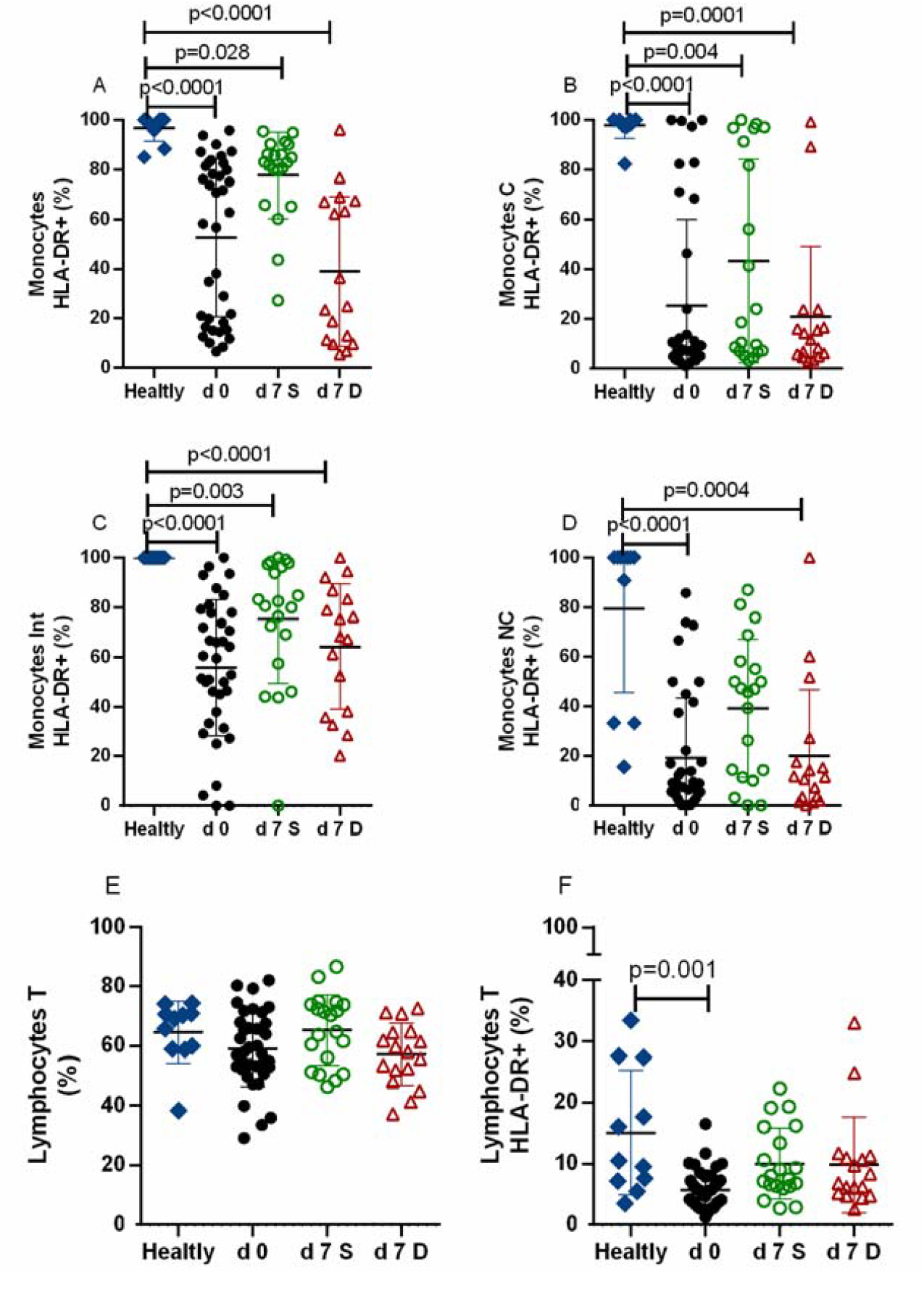
Percentage of HLA-DR+ leucocytes in Healthy donors and COVID-19 patients. Whole blood was phenotype as in methods. d0: Admission Day. d7: Seventh day in hospital. Blue diamonds show healthy donors. Red full circles show patients who death occurs during hospitalization not necessary at d0. Green empty circles show patients whose were discharge from hospital. Red empty triangles show patients whose fatal outcome occurs during hospitalization after 7 days of medical treatment. Monocytes Int: SSC^mid^FSC^mid^CD16^+^CD14^+^. Monocytes C: SSC^mid^FSC^mid^CD16^**-**^CD14^+^. Monocytes NC: SSC^mid^FSC^mid^CD16^+^CD14^**-**^. Results are expressed as mean±SD. Kruskal-Wallis and Dunn’s multiple comparisons test was calculated. Significant *p*<0.05.

Intermediate (Monocytes Int), and Non-Classical monocytes (Monocytes NC). We observed that the percentage of HLA-DR+ in each one of them are lower than in healthy donors, however, only in Monocytes NC the percentage of HLA-DR+ are similar between healthy patients and d7S patients (Fig 3D). The HLA-DR was also analyzed in T cells, and the percentage of this lymphocytes was similar between healthy and COVID-19 patients (Fig 3E), however, the percentage of T cells HLA-DR+ is lower in COVID-19 patients at the admission day d(0) than in healthy patients (Fig 3F).

The percentage of leucocytes CD39+, CD73+ and CD39+CD73+ were analyzed on leucocytes from healthy donors and COVID-19 patients. COVID-19 patients characteristically show a similar percentage of CD39 on monocytes and B-like cells than in the healthy donors. Our data show that the percentage CD39+ cells remain similar after seven days of treatment despite outcome (Fig 4A, 4G). In addition, and despite the final outcome (survive or deceased), the percentage of CD39 on T cells is higher in COVID-19 patients after the seventh day of hospitalization (Fig 4D), d7D patients show the highest percentage, however, was no statistical difference between d7S and d7D (Mann-Whitney test, p=0.167. Regarding the percentage of CD73 cells, we observed a higher percentage of CD73 on monocytes from COVID-19 patients than in healthy donors (Fig 4B), while B-like cells from COVID-19 patients only show a higher percentage of CD73 cells than in healthy donors (Fig 4H). In contrast, the percentage of leucocytes (monocytes, T and B cells) show a similar percentage of CD39+CD73+ cells from COVID-19 patients than in healthy donors (Fig 4C, 4F and 4J).

**Fig 4.**
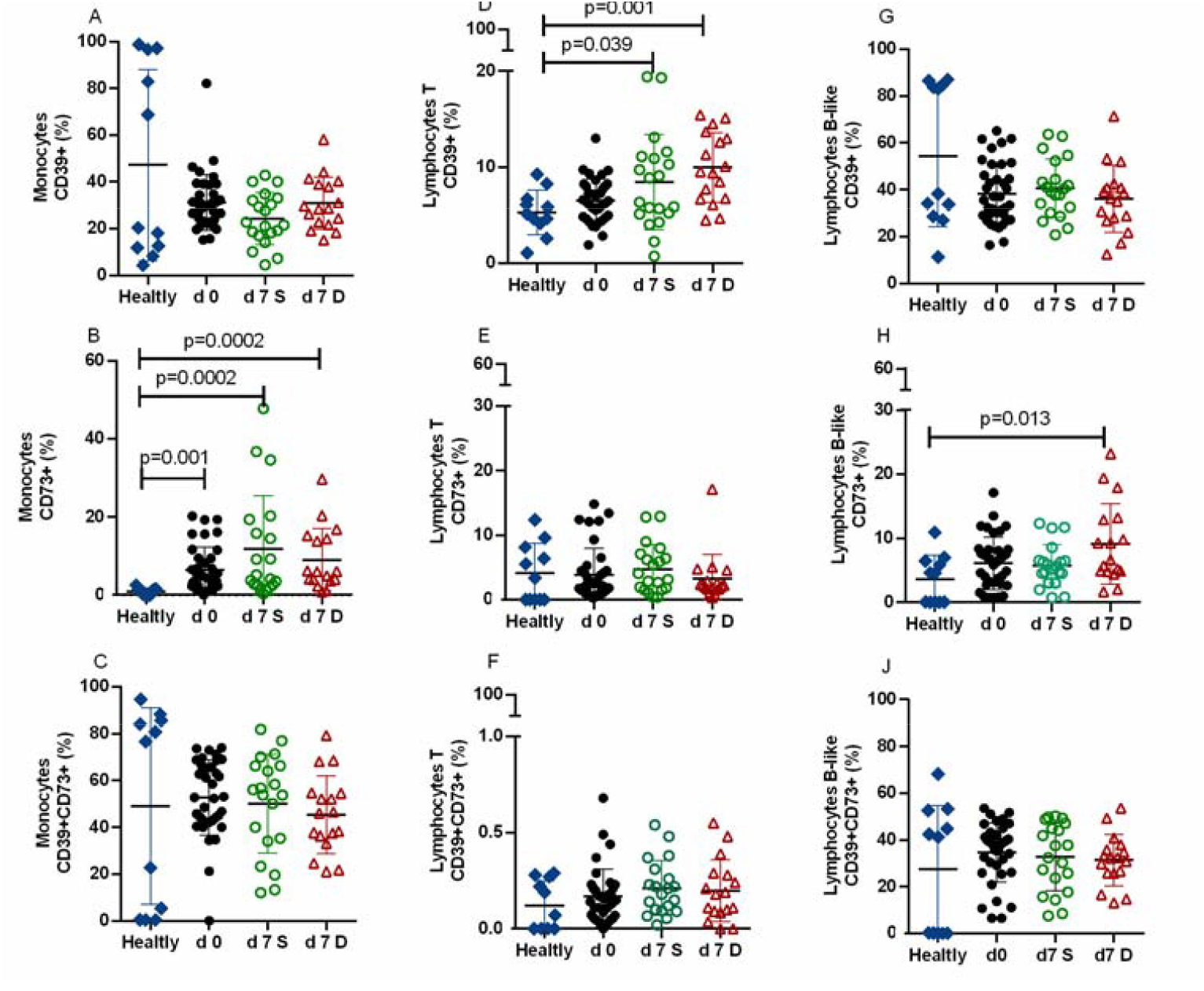
Percentage of CD39+, CD73+ and CD39+CD73+ leucocytes in COVID-19 patients. Whole blood was phenotype as in methods. d0: Admission day. d7: Seventh day in hospital. Blue diamonds show healthy donors. Red full circles show patients whom death occurs during hospitalization (not necessary at d0). Green circles show patients who were discharged alive from hospital. Red triangles show patients who fatal outcome occurs during hospitalization after 7 days of medical treatment. Results are expressed as mean±SD. Kruskal-Wallis and Dunn’s multiple comparisons test was calculated. Significant *p*<0.05.

## Discussion

Our study analyzed a cohort of patients in Mexico City from April 2020 to November 2020. Demographic, clinical, and laboratory data were recorded from the patients with COVID-19 from admission to discharge, with two possible outcomes: survival or death. Also peripheral leucocyte immunophenotype was performed to analyzed differential expression of inflammation related molecules as HLA-DR, CD39 and CD73 with prognostic potential in other hyperinflammatory states as sepsis or SIRS (Systemic Inflammatory Syndrome).

We test and/or verify which of these parameters could be predicting the fatal outcome of COVID-19 patients treated in a middle-income country with a high frequency of co-morbidities as in Mexico. ROC analysis were applied to variables that express difference between survivor and fatal outcome patients, and to know the ultimate cut-off value for each variable with useful ROC value.

It has been reported that some demographic or clinical factors may, in fact, be of good prognostic against COVID-19 [31]. Several international reports show that age up to 50 is a useful predictor of fatal outcomes [32]. In Mexico, Rizo-Téllez et al. report that 58.9±13.7 (media±SD) years old in non-survival COVID-19 patients, while survival patients were in 51.4±13.7 years old [33]. Concordantly, our study shows that non-survival patients were older and 59±14 years old, quite similar to the Rizo-Téllez report; however, after the multivariate logistical regression model, age alone in our study is not a useful predictor of fatal outcome. Also, our study did not find statistical differences in tachypnea between those who survived or died; however, there was a difference in oxygen saturation which was lower in those who died. Successful gas exchange depends partially on the respiratory rate; however, seems a higher respiratory rate was insufficient to get a successful gas exchange in Non-survivor patients.

On the other hand, with co-morbidities like obesity, some authors point out that it is a high risk factor for serious and fatal outcomes in COVID-19[34, 35]. Some reports show a clear association between severe COVID-19 and obesity, but this association is not so strong for fatal outcomes and obesity [36]. In contrast, we did not find a statistical difference in obesity or diabetes presence between survival and no survival patients. Still, hypertension and CRF were more frequent in those who died, which agrees with the recen reports for hispanic population pre-existing conditions assoc iated with fatal ourcomes of countries with a variety of income status[37].

Concerning laboratory determinations, the D-Dimer concentration has been proposed as a useful predictor of fatal outcomes [38], however, we did not observe a difference in the plasma concentration of D-Dimer between survival and non-survival groups. This could be due to differential endothelial dysfunction states related with both inflammatory and chronic diseases, since D-Dimer can be modified due to coagulopathy and endothelial activation[39].

As reported for sepsis and pancreatitis, our data showed SOFA score is a useful predictor of fatal outcomes in COVID-19. Also, lymphopenia is a variable consistently reported to help predict the progression of COVID-19 to more sever states[40]. Our study indicates that the NLR is useful as a predictor of fatal outcomes; moreover, the calculation of the pNLR is also helpful as a predictor of fatal outcome, reinforcing the observation made with the NLR. The NLR is a value that can be easily obtained and is calculated after performing a leukocyte count in peripheral blood. Such a count is within reach of most countries, and universal standardization exists to achieve it. Notably other demographic, clinical, or laboratory parameters are not always practical as sufficient predictors of the outcome, but the NLR can be consistently helpful even in patients with pre-existing co-morbidities. The higher the NLR value, the greater probability of a fatal outcome in patients with COVID-19; the higher value may result from the increase in neutrophils and/or the decrease in lymphocytes in the patient with COVID-19. We think that the chemokine-dependent neutrophilia could be a factor that explains the massive mobilization of these cells in peripheral blood. However, despite their high protective function, phagocytic or microbicidal capacity could be limited in neutrophils, and neutrophilia could be a mechanism to compensate for the lack of function. Likewise, lymphopenia can result from the redistribution of lymphocytes to elaborated tissue and the subsequent damage of the tissue to limit viral replication in COVID-19. This condition is favored as toxic for the lymphocyte population, especially for cytotoxic lymphocytes. More studies must be carried out to determine the cause of the increase in the value of the NLR and to be able to influence the treatment that improves the patients’ conditions with the use of the NLR as a monitor.

Regarding cytokines and chemokines, we show that IL-6, IL-10, CXCL10 (IP-10), and CXCL11 are higher in COVID-19 patients than in healthy, indicating that SARS-CoV-2 infection evokes a state of hyperinflammation with a composite component, pro, and anti-inflammatory. We could speculate that the plasma elevation of specific chemokines such as CXCL8, CCL4, CCL20, and CCL2 in those who die, that the inflammatory response -mainly affects molecules with chemoattractant function. However, there were no differences between those who survived and those who died. This is accordance with other soluble candidates as calcium in sera, that in initial studies seem to have prognostic potential, but when transferred to other populations than Chinese it failed as a predictor of severity. This could be due to the different cut-off points used for the analytes instead of the actual plasma levels[41].

The hypercytokinemia reported in COVID-19 includes IL-6, TNF-α, and IP-10. Our study shows that patients with a fatal outcome consistently express higher plasma concentrations of inflammatory mediators; however, when performing the ROC curve analysis, only IL-6 was a good predictor for a fatal outcome, regardless of the way in which it was expressed. As we observed for the NLR value, the high concentration of IL-6 could suggest that COVID-19 is not necessarily a condition with general hypercytokinemia, but rather a specific dysregulation of the response of various tissues through IL-6. This cytokine has a broad pleiotropic response and synergizes several responses to other cytokines. This is in accordance with the improved clinical conditions and reduced mortality reported with anti-IL-6R treatments in patients with severe COVID-19 [42].

Finally, we analyzed the percentage of monocytes and T lymphocytes that express HLA-DR. Our group previously reported that the percentage of monocytes HLA-DR+ is diminished in septic patients and is associated with poor prognosis [20]. Here we show that the percentage of HLA-DR+ in monocytes from COVID-19 patients is lower than in healthy subjects (Fig 3A), also d7D patients expressed a lower percentage of HLA-DR+ monocytes than in d7S patients (Mann-Whitney test, p<0.0001), this was closer to reach statistical difference in Non-classical monocytes (Mann-Whitney test, p=0.057). However, after ROC curve analysis it does not reach a high sensitivity or specificity to be a good predictor of fatal outcome in COVID-19 patients. More studies are necessary to know if HLA-DR expression on monocytes could be useful to predict fatal outcome in COVID-19 patients. Also, we explored the expression of other surface markers (CD39 and CD73) that could limit the inflammatory response in COVID-19. The percentage of CD39 on T cells was higher in COVID-19 patients after seven days of treatment; this was not the case for monocytes and B-like cells, indicating that some anti-inflammatory molecules are differentially expressed in peripheral leucocytes, however, we do not know if leucocytes in tissue expressed this differential condition, the expression of CD39/CD73 could regulate the activation of leucocytes *in situ* and limit the inflammatory response in COVID-19 more effectively. Our result shows a higher percentage of monocytes and B-like CD73+ cells than healthy donors, indicating that the expression of CD73 could be more important to regulate anti-inflammatory response in monocyte and B cells than in T lymphocytes. Our result about CD73 agrees with the report of Díaz García et al. [43], but in contrast to Ahmadi et al. [26]. Ahmadi et al found a lower expression of CD73 in CD4, CD8 T lymphocytes, while we did not observed a difference (Fig 4E), also, Ahmadi reported a lower expression of CD73 in monocytes and B lymphocytes, while we observed a higher percentage which not always reach statistical difference, this difference could be explained because we include the outcome in the analysis [26]. Finally, the expression of both CD39 and CD73 on the same cell was similar in COVID-19 patients and healthy donors (Fig 4C, 4F, and 4J), suggesting that monocytes and B and T cells have the same condition to regulate inflammation by CD39/CD73 molecules. Our result shows that single CD39 or CD73 expression on leucocytes have some difference in peripheral blood, however, double expression of CD39 and CD73 on cells is similar in COVID-19 patients and healthy donors, suggesting that this anti-inflammatory mechanism is working in COVID-19, taking together these results suggest that SARS-CoV-2 infection could dissociate the expression of CD39 and CD73, increasing the single cells and limiting the anti-inflammatory functionality of the pair CD39/CD73. More studies are necessary to clarify this possibility.

The pandemic came as a new medical challenge, and we had limited medical resources that restrain our capacity to offer the attention of COVID-19 patients in hospital. Our study included a high number of patients with poor outcomes, whichg focus and limited our study to a specific group of patients with high severe conditions, which prevented having a common moderate or mild covid group to compare and establish if our observations are related to the disease and/or severity. A disbalance number of observations for some variables is another limitation. As a relative limitation is that it is a single-center study. Also, we assume that there has been no re-infection or asymptomatic COVID-19 cases in COVID-19 patients or healthy donors, respectively. The interpretation of the results should consider these limitations.

## Data Availability

All data produced in the present study are available upon reasonable request to the authors

## Acknowledgements

The authors thank the staff at the Hospital de Especialidades, Centro Médico Nacional Siglo XXI “Dr. Bernardo Sepúlveda”.

(Hidden peer review process)

