## Supplementary figures and images for "Potential biomarkers for fatal outcome prognosis in a cohort of hospitalized COVID-19 patients with pre-existing co-morbidities"

### Supplemental Figure 1

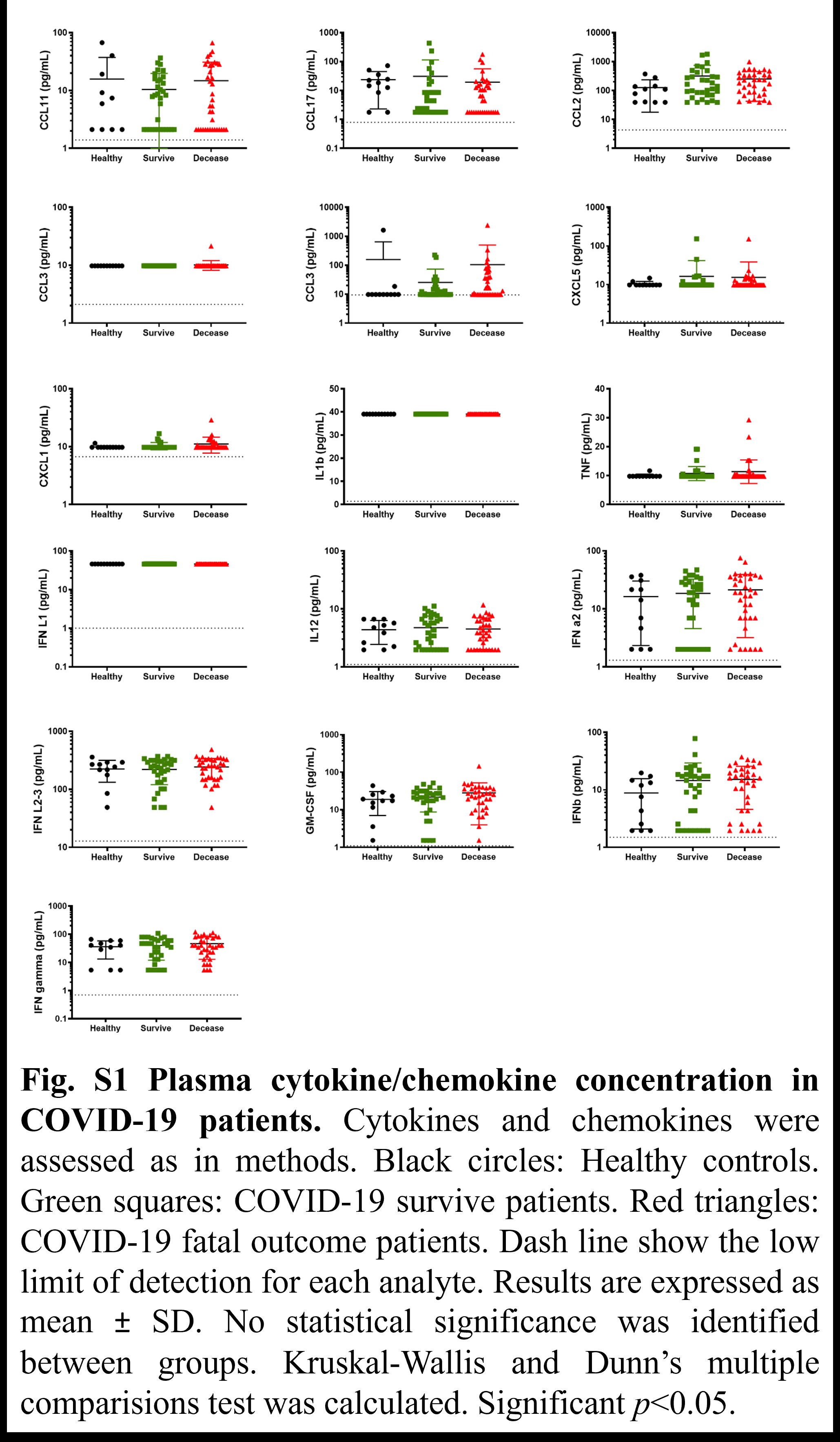

### Supplemental Figure 2

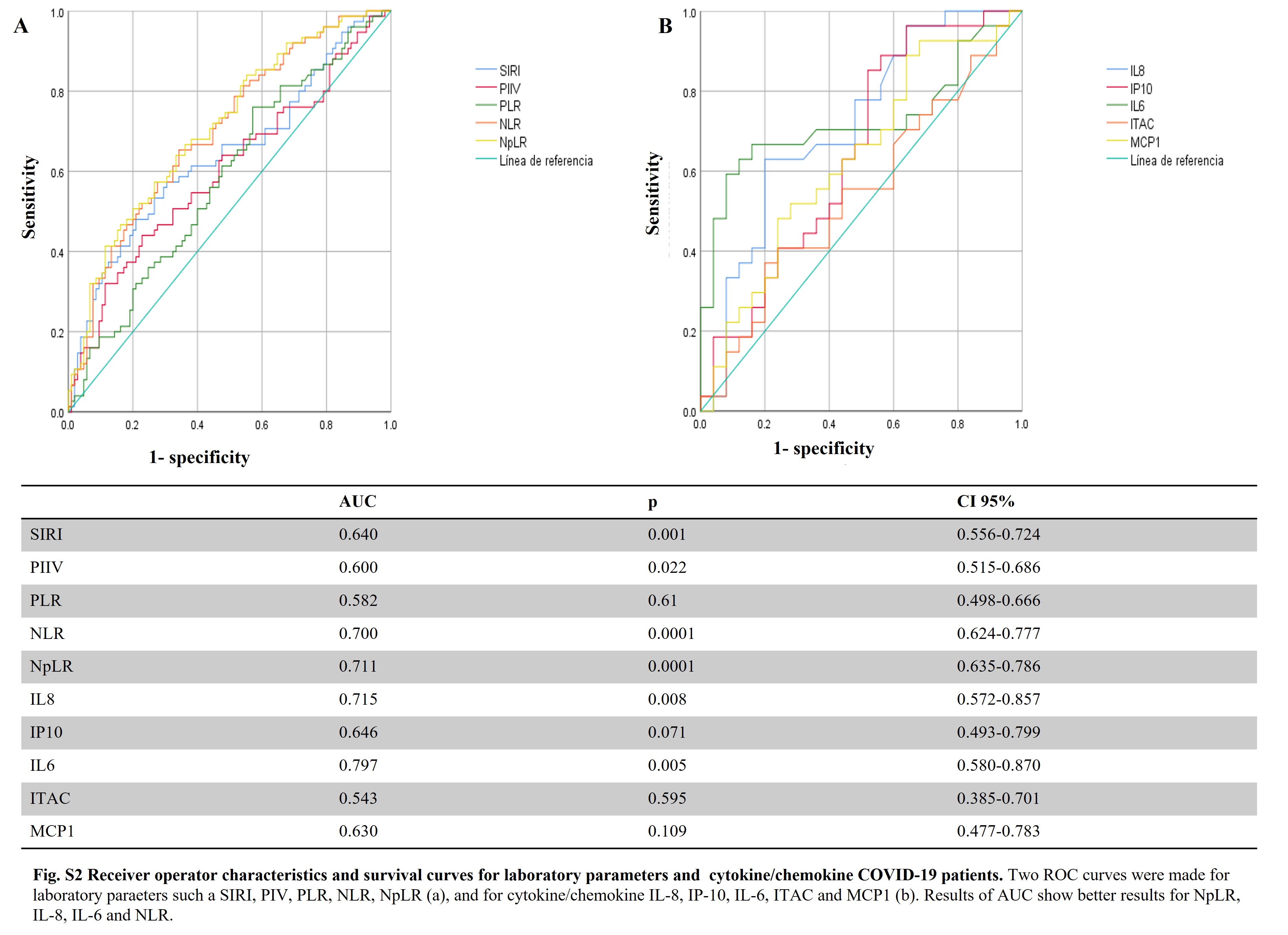
